# Impaired functional connectivity in patients with psychosis and visual hallucinations

**DOI:** 10.1101/2022.05.06.22274666

**Authors:** Marouska van Ommen, Azzurra Invernizzi, Remco J. Renken, Richard Bruggeman, Frans W. Cornelissen, Teus van Laar

**Affiliations:** Department of Neurology, University Medical Center Groningen, University of Groningen, Groningen, The Netherlands; Laboratory for Experimental Ophthalmology, University Medical Center Groningen, University of Groningen, Groningen, The Netherlands; Cognitive Neuroscience Center, Department of Biomedical Sciences of Cells & Systems, University Medical Center Groningen, Groningen, the Netherlands; Department of Psychiatry, University Medical Center Groningen, University of Groningen, Rob Giel Research Center, Groningen, The Netherlands; Department of Clinical Neuropsychology, University of Groningen, Groningen, The Netherlands

**Keywords:** visual hallucinations, schizophrenia, resting-state functional MRI, functional connectivity, eigenvector centrality mapping

## Abstract

**Background:** more than one-third of patients with psychosis experience visual hallucinations, but the underlying pathomechanism remains largely unknown. Although schizophrenia is related to altered brain functional connectivity, it is unknown how this could predispose patients to experience visual hallucinations. Previous work suggested that this predisposition is caused by alterations in vision-related networks, including the Visual Network, possibly with a specific focus on the Ventral Attention Network ^1^. This network responds to salient stimuli from the Visual Network and operates as a switch between the internally-focused Default Mode Network and the outside-world-focused Dorsal Attention Network.

**Methods:** in this case control study we investigated the role of these networks in three groups: 14 participants with a psychotic disorder and visual hallucinations, 15 participants with a psychotic disorder without visual hallucinations, and 16 healthy controls. All patients underwent resting state functional Magnetic Resonance Imaging after which we determined the intra- and inter-network functional connectivity of these networks in all participants. We also used fast Eigenvector Centrality Mapping to determine the most central regions, i.e. the most functionally communicating regions, within these networks.

**Results:** compared to healthy controls, patients with visual hallucinations had lower functional connectivity, both intra-network and inter-network, in all vision-related networks. This decrease was most prominent for the Ventral Attention Network and the Dorsal Attention Network for intra-network functional connectivity. Moreover, Eigenvector Centrality Mapping showed a severe decrease in functional communication within the Visual Network in the right intracalcarine sulcus, with a simultaneous increase in functional communication in the lateral part of the left middle occipital gyrus, a region involved in object recognition. The results of patients without hallucinations were generally in between patients with visual hallucinations and healthy controls.

**Discussion:** our study shows that widespread dysconnectivity of predominantly vision-related functional networks may predispose patients with psychosis to generate visual hallucinations. These results are in line with previous models of hallucinations in psychosis which suggested that the processing deficits in the Visual Network may cause or exacerbate inadequate co-functioning and switching between the Default Mode Network and the Dorsal Attentional Network, possibly due to impaired Ventral Attention Network functioning. In combination with impaired attending of visual signals by the Dorsal Attentional Network, this may lead to inappropriate saliency processing and wrongly attributing an external origin to internally generated events and, consequently, to visual hallucinations. The often complex nature of psychotic visual hallucinations may be explained by the more central role of object processing regions.

## Introduction

Psychotic disorders are characterized by the occurrence of hallucinations, delusions, disorganized speech, disorganized or catatonic behavior, and negative symptoms such as apathy ^2^. The most common psychotic disorder is schizophrenia. As early as the 19^th^ century, schizophrenia -– which was still called amentia or dementia praecox – was thought to be caused by altered brain connectivity ^3^. Recent studies have supported this hypothesis; compared to healthy controls, schizophrenia regions and networks throughout the brain appear to be to less functionally connected to each other, and nodes within networks less hierarchically organized ^4,5^. However, it remains largely unknown how visual hallucinations (VH), which occur in 37% of patients with psychosis, are related to altered brain connectivity ^6^. We therefore analyzed fMRI data acquired during resting state and determined the functional connectivity and most central brain regions (‘hubs’) in the vision-related functional networks in two groups of participants with a psychotic disorder: those who have experienced VH and those who have not.

VH in psychosis are usually complex, and include images of people and animals ^7,8^. Shine et al. hypothesized that these complex VHs are caused by dysfunction both within and between the vision-related networks ^1^. These functional networks encompass the Visual Network (VIS), the Default Mode Network (DMN), the Dorsal Attention Network (DAN) and the Ventral Attention Network (VAN). The VAN, including the anterior insula and ventral frontal cortex, responds to salient stimuli as captured by the VIS. The VAN switches processing between the internally-focused DMN and the outside world-focused DAN (see Fig. 1A). The DAN, including the frontal eye fields and dorsolateral prefrontal cortex, primes visual objects in the external world by focusing on them. Contrarily, the DMN mainly consists of medial cortical regions and is active when the brain is at rest or internally focused, such as during memory retrieval and when envisioning the future ^9^. Both the DMN and DAN interact with the VIS to elucidate the stimulus’ content ^1^. Dysfunctional connectivity, both within and between these functional networks, may lead to an erroneous interpretation of ambiguous signals, and thus to VH ^1^. Moreover, the VAN would play a central role in the generation of VH in schizophrenia by constantly generating superfluous neural responses to both externally and internally evoked signals. Additional aspects contributing to VH could include impaired processing in the VIS, the DAN’s inability to appropriately prime stimuli, and hyperactivity and hyperconnectivity of the DMN. VH in schizophrenia are also related to higher white matter connectivity between the hippocampus (DMN) and visual areas ^10^. These studies therefore suggest that altered functional connectivity both within and between vision-related networks predisposes some patients with psychosis to generate VH.

**Fig. 1).**
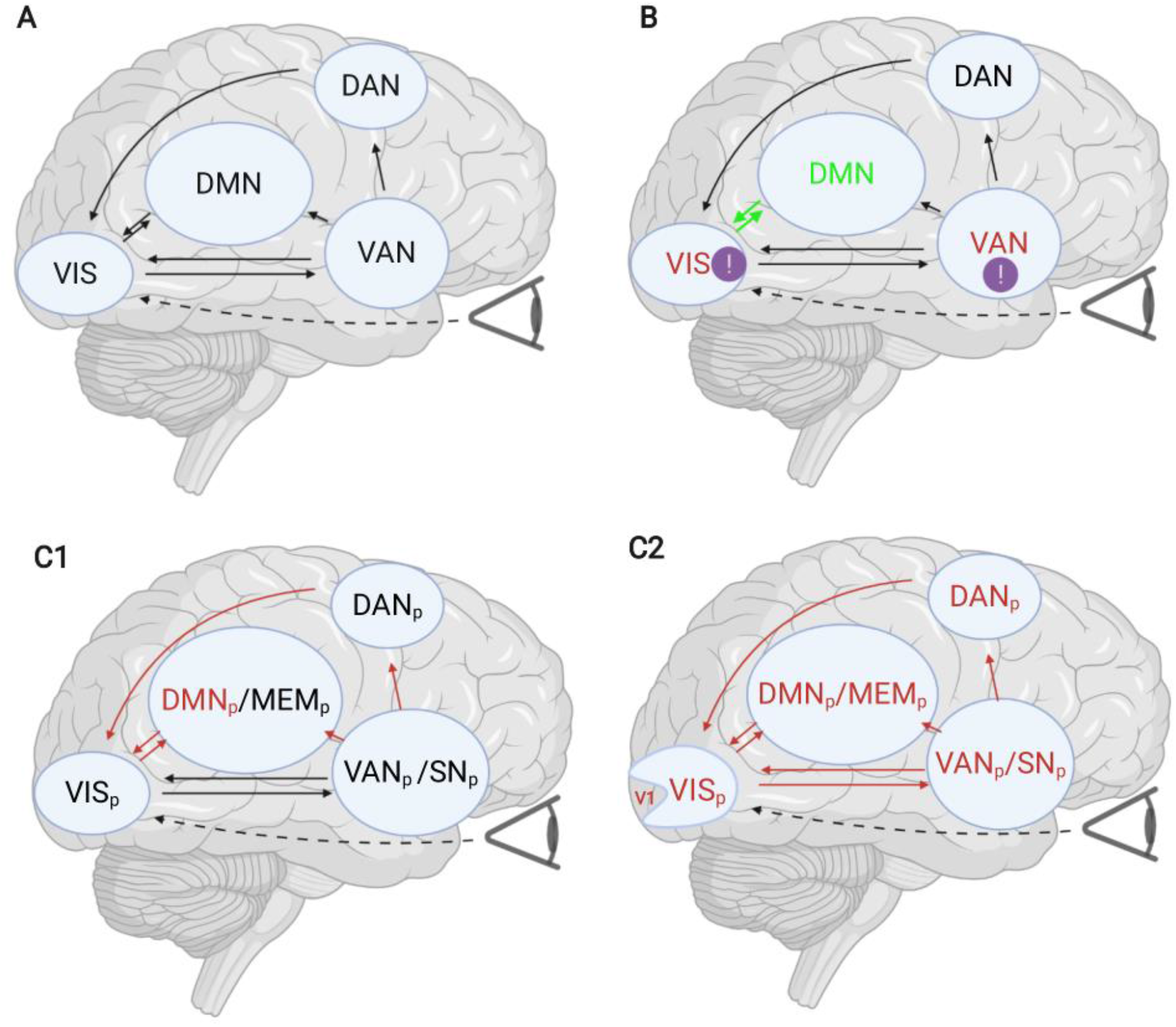
visualization of visual processing, our hypothesis and a summary of the results. **A** The networks involved with normal visual processing, based on ^1^, with the author’s permission. The networks involved with visual processing are the visual system (VIS), the Default Mode Network (DMN), the Dorsal Attention Network (DAN) and the Ventral Attention Network (VAN). The VAN responds to salient stimuli as captured by the VIS: it switches activation away from the inner-focused DMN to the outer-focused DAN. The DAN primes visual stimuli in the external world. The DAN and DMN themselves also interact with VIS to elucidate the content of the stimuli. **B** Our hypotheses. Mostly based on ^1^, with the author’s permission, but also on ^10^. Altered functional connectivity both within and between vision-related networks would predispose some patients with psychosis to generate VH, via erroneous interpretation (having an origin in the external world) of ambiguous signals. The VAN is thought to play a key role in psychotic VH generation by constantly generating superfluous neural responses to both externally and internally evoked signals. Moreover, impaired processing in the VIS, the DAN’s inability to appropriately prime stimuli, and hyperactivity and hyperconnectivity of the DMN would contribute to VH. VH in schizophrenia are also related to higher white matter connectivity between the hippocampus (DMN) and visual areas (10). Thus, previous studies suggest altered functional connectivity both within and between vision-related networks, which predisposes some patients with psychosis to generate VH. Green letters: higher intra-network functional connectivity. Red letters: lower intra-network functional connectivity. Green arrows: higher intra-network functional connectivity. Red arrows: lower inter-network functional connectivity. Exclamation mark: important role. **C1** Results of comparisons between PSVH- (patients with a psychotic disorder without visual hallucinations) and HC (healthy controls). Green letters: higher intra-network functional connectivity. Red letters: lower intra-network functional connectivity. Green arrows: higher intra-network functional connectivity. Red arrows: lower inter-network functional connectivity. PSVH-have lower intra-network FC compared to HC for the DMN_p_, and lower inter-network FC than for VIS_p_-DAN_p_, VIS_p_-DMN_p_, VAN_p_-DAN_p_, SN_p_-DAN_p_, VAN_p_-DMN_p_, DAN_p_-DMN_p_. PSVH- and HC do not differ regarding the distribution of eigenvector values. **C2** Results of comparisons between PSVH+ (patients with a psychotic disorder with visual hallucinations) and HC (healthy controls without visual hallucinations). PSVH+ have lower intra-network FC than HC for VIS_p_, VAN_p_, SN_p_, DAN_p_ and DMN_p_, and lower inter-network FC compared to HC for the following combinations: VIS_p_-SN_p_, VIS_p_-DAN_p_, VIS_p_-DMN_p_, SN_p_-VAN_p_, SN_p_-DAN_p_, SN_p_-DMN_p_, VAN_p_-DAN_p_, VAN_p_-DMN_p_, and the DAN_p_-DMN_p_. ECM: compared to HC, PSVH+ have lower ECM values for the right ICalc than HC, lying closely to H0. PSVH+ have lower ECM values for the left MTGo than HC, and a strong trend towards higher left MOGl. Green letters: higher intra-network functional connectivity. Red letters: lower intra-network functional connectivity. Green arrows: higher intra-network functional connectivity. Red arrows: lower inter-network functional connectivity. V1: lack of communication.

Functional connectivity within and between networks (intra-network FC and inter-network FC) can be assessed with resting state functional Magnetic Resonance Imaging (rs-fMRI) ^11^. Rs-fMRI is task-free, thereby placing minimal demands on participants and circumventing task-related confounding factors ^12^. It is therefore a useful method for clinical populations, such as patients with psychosis ^13,14^. More specific knowledge about what brain mechanisms play a role in the generation of VH could be given by determining the most important regions within networks. On this smaller, sub-network scale, the functional organization of the brain can also be depicted as a graph consisting of multiple ‘nodes’ (brain regions) and ‘edges’ (connections) ^15^.

From these parameters the relative importance of a node within the overall architecture of a network can be calculated. A node occupying a central position in the overall organization of a network is called a ‘hub’. If we knew more about the hubs within networks in patients with VH, this could lead to more precise knowledge about the occurrence of VH.

In the present study, we first compared whole-brain FC in three groups: patients with a psychotic disorder with VH, patients with a psychotic disorder but without VH, and healthy controls. Second, we compared the intra-network and inter-network FC of vision-related networks in these three groups. Third, we identified the hubs in these networks for each group and compared them between groups by using Eigenvector centrality mapping. In these studies we addressed five hypotheses: 1) that patients with psychosis (both with and without VH) have lower overall FC than controls; 2) that patients with VH have lower FC intra-VIS, lower intra-VAN, and higher FC intra-DMN than controls; 3) that patients with VH have higher FC between VIS-DMN; 4) that patients with VH show more (or more prominent) intra-network and/or inter-network FC changes or hubs within the VAN; and finally, 5) that patients with VH have more or more central hubs in higher visual areas compared to patients without VH. Our hypotheses are visualized in Fig. 1B.

## Materials and methods

### Participants

This case control study took place at the NeuroImaging Center at the University Medical Center Groningen from 08-09-2015 to 30-03-2018. It is part of the INZICHT trial, which has the goal of gaining insight into VH in psychosis. It involves a cognitive component (INZICHT1, https://www.trialregister.nl/trial/4858), and an fMRI component (INZICHT2; including the present study, https://www.trialregister.nl/trial/6685). Participants in INZICHT2 were recruited via: 1) INZICHT 1, 2) the GROUP study, a multi-site longitudinal observational study focused on gene-environment interaction ^16^, 3) the Department of Psychotic Disorders, University Medical Centre Groningen (UMCG), 4) Lentis Center for Mental Health Groningen and Winschoten; 5) Anoiksis, an association for patients with psychotic disorders.

The following inclusion criteria were applied: 1) age between 18-55; 2) fluent speaker of Dutch; 3) able to give informed consent. In addition, participants also had to meet the DSM-IV-TR criteria (or the DSM 5 equivalent) for schizophrenia, schizophreniform disorder, schizoaffective disorder or psychotic disorder Not Otherwise Specified (NOS) ^17^. Included patients fell into two groups: those who experienced VH more than a couple of times in the last month (patient group, PSVH+) or almost never had VH (patient group PSVH-). All PSVH-participants had never experienced VH, with the exception of one who experienced VH once around 7 years ago, an another who experienced VH twice 8 years ago. In case of psychiatric comorbidity, the psychotic disorder had to be predominant. Moreover, the VH had to be related to the primary psychotic disorder. Their own psychiatrist evaluated the last two conditions. Exclusion criteria were: 1) psychiatric disorders: mental retardation, amnesia, delirium, current substance dependence (excluding nicotine and caffeine), dissociative disorders and borderline personality disorder; 2) neurological disorders: dementia, degenerative, demyelinating or inflammatory diseases of the central nervous system, and other non-congenital anatomical cerebral abnormalities such as tumors and infarcts, epilepsy, congenital brain injury, brain surgery, current mild traumatic brain injury or a medical history of more severe traumatic brain injury; 3) visual acuity <50% (assessed by a chart with sentences at a reading distance); 4) visual field defects (Donders technique); 5) cognitive impairment (Mini-Mental State Examination (MMSE) score <26 ^18^; and 6) fMRI incompatibility. Furthermore, healthy controls (HC) were excluded if they had experienced a psychotic episode or VH, or if a first degree family member had a history of psychosis. Participants received a 50 euro coupon for participating in INZICHT2.

The ethics board of the University Medical Center Groningen (UMCG) approved the study protocol. All participants provided written informed consent. The study followed the tenets of the Declaration of Helsinki.

### Symptom assessment

Trained researchers interviewed participants about psychotic symptoms using the Dutch version of the Questionnaire for Psychotic Experiences (QPE; http://qpeinterview.com/home/, ^19^) and the validated Positive and Negative Syndrome Scale (PANSS; ^20^).

### Visual evaluation

The following vision aspects were assessed at the Ophthalmology Department (UMCG): visual acuity (digital Snellen chart), contrast sensitivity (GECKO chart) and visual fields (computerized; Humphrey Field Analyzer, 24-2 SITA; Carl Zeiss Meditec, AG, Jena, Germany). An ophthalmologist assessed the visual fields results.

### MRI and fMRI data acquisition

Participants underwent a 10-minutes fMRI scan, during which they were instructed to keep their eyes closed, relax and think of nothing in particular, but not go to sleep. The lights in the scanner room were dimmed during scanning. Data was collected using a 3T Philips Magnetic Resonance system with a standard 64-channel SENSE head coil (Intera, Philips Medical Systems, Best, the Netherlands). Echo-planar images (EPI) were acquired, anterior-posterior, with the following parameters: TR 2 s, TE 30 ms, voxel size 3 mm isotropic, flip angle 90° (FOV 192×117×192), 39 slices per volume, without slice gap. A T1 weighted scan was acquired for anatomical reference (160 slices, voxel size 1 mm isotropic, FOV 256×160×224 mm).

Immediately after scanning, participants were asked if they had experienced hallucinations in any sensory domain during scanning. If so, we interviewed them about hallucination characteristics.

### Data analyses

Image preprocessing, FC analysis and statistical analyses were performed using SPM12 (Wellcome Department of Imaging Neuroscience, London, UK), fastECM toolbox ^21^, MarsBaR ROI toolbox (http://marsbar.sourceforge.net/) and customized scripts, implemented in Matlab 2016b (The Mathworks Inc., Natick, Massachusetts).

### Image preprocessing

For each subject, the structural MR image was co-registered and normalized to the Montreal Neurological Institute (MNI) template and segmented in order to obtain white matter (WM), grey matter (GM) and cerebrospinal fluid (CSF) probability maps in MNI space.

FMRI data were spatially realigned, co-registered to the MNI-152 EPI template, and subsequently normalized utilizing the segmentation option for EPI images in SPM. All normalized data were denoised using ICA-AROMA ^22^ and resampled to a 3 mm isotropic voxel resolution. Additionally, spatial smoothing was applied (8 mm) to fMRI. No global signal regression was applied.

### Pre-whitening

To facilitate statistical inference, data were “pre-whitened” by removing the estimated autocorrelation structure in a two-step GLM procedure ^23,24^. In the first step, the raw data were filtered against the 6 motion parameters (3 translations and 3 rotations). Using the resulting residuals, the autocorrelation structures present in the data were estimated using an Auto-Regressive model of order 1 (AR(1)) and then removed from the raw data. Next, the realignment parameters, white matter (WM) and cerebrospinal fluid (CSF) signals were removed as confounders on the whitened data.

### ROI and functional network definitions

Based on the Power atlas coordinates ^25^, 11 functional networks and the associated 232 regions of interest (ROI) were defined (5 mm radius) using the MarsBar ROI toolbox for SPM12 ^26^. For each ROI, a time series was extracted by averaging across voxels per time point. Because the network definitions in Shine and Power use similar terminology, we indicated the Power definition by the subscript “_p_” when applicable. In all other cases, we referred to the Shine nomenclature. Our hypothesis involved the DAN, VAN, VIS and the DMN, as defined by Shine et al. ^1^. In the Power atlas, this resulted in a combination of the Memory Retrieval Network (MEM_p_) and DMN_p_, thus creating the equivalent of the DMN, and in a combination of the Salience Network (SN_p_) and the VAN_p_, thus creating the VAN. Connectivity results for the other networks are included in the Suppl. Fig. 2 and 3. Due to their related function, the Sensory/somatomotor Hand and Mouth networks were combined into one network. We excluded the Cerebellar and Uncertain networks.

### Functional connectivity analysis

For each subject, the pairwise temporal Pearson correlation between ROIs was calculated and a Fisher’s z-transformed was applied. The ROI’s z-values (hereafter: FC values) were averaged across subjects. Then, the median group FC-values were used for the whole-brain analysis. Moreover, we computed the median group FC-value per single Power’s NWs. We used these values to perform the intra- and inter-network analyses.

For whole brain, intra-network and inter-network analyses, the median group FC values were compared between groups using a family-wise error-corrected (FWE) permutation test. The subject’s permuted group labels were repeated 10,000 times; p≤ 0.05 was considered statistically significant. To identify the alterations to which VH related most often, effect sizes (ES) for both intranetwork and inter-network FC were ranked.

### Fast Eigenvector Centrality Mapping

To identify the most important hubs of the predefined networks, fast ECM ^21^ was performed on the defined ROI time course data per subject. The ECM method builds on the concept of node centrality, which characterizes functional networks active over time and attributes a voxel-wise centrality value to each ROI. Such a value is strictly dependent on the sum of centrality properties of the directly neighboring nodes within a functional network. In the fast ECM toolbox ^21^, ECM is estimated from the adjacency matrix, which contains the pairwise correlation between the ROIs. To obtain a real-valued ECM value, we added +1 to the values in the adjacency matrix. Several ECM values can be attributed to a given node, but only the eigenvector with the highest eigenvalue (EV) is used for further analyses. The highest EV values were averaged across subjects at group level. Based on these values, influential ROIs, i.e. the hubs ^27–29^, can then be identified. For the FC analyses, we identified these hubs only for our NW of interest (VIS_p_, VAN_p_, SN_p_, DAN_p_, DMN_p_, MEM_p_), based on Power’s definition. Per group, only the ROIs with the 5% highest ECM coefficients (the hubs) were considered in subsequent analysis (see Suppl. Fig. 1 for the highest 10%).

To address group differences for the most influential hubs, mean group ECM coefficients were compared between groups. FWE correction was applied for the number of group level comparisons, but not for the total number of ROIs analyzed. Here as well, permuted labels were repeated 10,000 times; p≤ 0.05 was considered statistically significant. In addition, a proxy distribution for the null hypothesis (H0) was obtained by generating a surrogate BOLD time series 1000 times using the iterative amplitude-adjusted Fourier transform method (iAAFT) ^30,31^. In this way, correlations between ROIs were removed; the null distribution represents the amount of centrality obtained when no functional communication is present in the brain. Note that the null distribution of the ECM is not centered at zero, as ECM values are forced to be positive real-valued. To define the confidence intervals of each ECM value estimated per ROI, a bootstrap technique (across time-point) was used at group level in parallel to resample the filtered fMRI data. To support visualization, a Gaussian distribution was fitted to both bootstrap and surrogate distributions.

### Data and code availability

The raw data and code belonging to this study are available from the corresponding author upon reasonable request.

## Results

We initially included 48 participants: 15 PSVH+, 16 PSVH- and 17 HC. One PSVH+ participant was excluded because the scan was not saved correctly, one PSVH-was excluded due to excessive motion (>3mm) and one HC was excluded due to a structural brain abnormality. This led to 14 PSVH+, 15 PSVH- and 16 HC participants being included in the further analyses. Table 1 shows the demographic and illness characteristics.

**Table 1:** demographic and illness characteristics.

**A**
|  | **Patients**<br>**(n=29)**<br>**mean (SD)** | **HC**<br>**(n=16)**<br>**mean (SD)** | **Test score**<br>**(df)** | **p-value** |
| --- | --- | --- | --- | --- |
| Age (y) | 36.0 (9.5) | 32.4 (11.2) | 182.00 | .24 |
| Gender (n, (%)) |  |  | 18 (1) | .74 |
| Males | 20 (69.0) | 12 (75.0) |  |  |
| Females | 9 (31.0) | 4 (25.0) |  |  |
| Education* | 6.4 (1.5) | 7.5 (0.7) | 335.0 | .01 |
| MMSE<sup>#</sup> | 28.8 (1.1) | 29.7 (0.5) | 336.5 | .01 |
| Visual acuity ODS<sup>^</sup> | 1.2 (0.2) | 1.3 (0.2) | 279.0 | .23 |
| Visual contrast ODS<sup>-</sup> | 15.0 (1.2) | 15.4 (0.9) | 255.0 | .28 |
| Diagnosis (n, (%)) |  |  |  |  |
| Schizophrenia | 16 (55.2) |  |  |  |
| Schizoaffective disorder | 6 (20.7) |  |  |  |
| Psychosis NOS | 7 (24.1) |  |  |  |
| Disease duration (y) | 13.1 (11.2) |  |  |  |
| PANSS<sup>&</sup> |  |  | 18.4 (3,41) | .00 |
| tot pos | 16.7 (5.4) | 7.6 (0.7) | 43.5 (1) | .00 |
| tot neg | 14.9 (4.6) | 7.9 (1.6) | 33.8 (1) | .00 |
| tot gen | 34.9 (9.0) | 18.3 (1.5) | 54.2 (1) | .00 |
| tot | 66.5 (17.0) | 33.8 (2.4) | 57.8 (1) | .00 |
| QPE<sup>+</sup> |  |  | 5.1 (4,40) | .00 |
| severity VH | 5.8 (6.8) | 0 (0) | 11.5 (1) | .00 |
| severity AH | 9.6 (10.8) | 0 (0) | 12.3 (1) | .00 |
| severity all H | 16.4 (15.5) | 0 (0) | 17.8 (1) | .00 |
| severity delusions | 4.9 (5.5) | 0 (0) | 12.6 (1) | .00 |
| Use antipsychotics (n, (%)) |  |  |  |  |
| Yes | 23 (79.3) |  |  |  |
| No | 6 (20.7) |  |  |  |
| Type antipsychotics (n, (%)) |  |  |  |  |
| 1 typical | 1 (4.3) |  |  |  |
| 1 atypical | 15 (65.2) |  |  |  |
| Multiple | 7 (30.4) |  |  |  |
| Other medication<sup>l</sup> (n, (%)) |  |  | 2.35 | .33 |
| Yes, 1 | 12 (41.4) | 3 (20.0) |  |  |
| No | 11 (37.9) | 9 (60.0) |  |  |
| Multiple | 6 (20.7) | 3 (20.0) |  |  |

B
|  | HC<br>(n=16)<br>mean (SD) | PSVH+<br>(n=14)<br>mean (SD) | PSVH-<br>(n=15)<br>mean (SD) | Test score<br>(df) | p-value |
| --- | --- | --- | --- | --- | --- |
| Age (y) | 32.4 (11.2) | 34.9 (8.1) | 37.0 (10.7) | 1.58 (2) | .45 |
| Gender (n, (%)) |  |  |  | 1.9 (FE) | .42 |
| Males | 12 (75.0) | 8 (57.1) | 12 (80.0) |  |  |
| Females | 4 (25.0) | 6 (42.9) | 3 (20.0) |  |  |
| Education* | 7.5 (0.7) | 5.9 (1.7) | 6.9 (1.2) | 9.0 (2) | .01<sup>B</sup> |
| MMSE<sup>#</sup> | 29.7 (0.5) | 28.9 (1.1) | 28.9 (1.1) | 7.1 (2) | .03 |
| Visual acuity ODS<sup>^</sup> | 1.3 (0.2) | 1.1 (0.2) | 1.2 (0.2) | 4.42 (2) | .11 |
| Visual contrast ODS<sup>~</sup> | 15.4 (0.9) | 14.9 (1.4) | 15.2 (1.0) | 1.17 (2) | .56 |
| Diagnosis (n, (%)) |  |  |  | 2.1 | .37 |
| Schizophrenia |  | 7 (50.0) | 9 (60.0) |  |  |
| Schizoaffective disorder |  | 2 (14.3) | 4 (26.7) |  |  |
| Psychosis NOS |  | 5 (35.7) | 2 (13.3) |  |  |
| Disease duration (y) |  | 11.1 (9.0) | 15.1 (12.6) | 126.0 | .38 |
| PANSS<sup>&</sup> |  |  |  | 9.8 (6,80) | .00 |
| tot pos | 7.6 (0.7) | 19.3 (4.5) | 14.3 (5.2) | 33.0 (2) | .00<sup>A,B,C</sup> |
| tot neg | 7.9 (1.6) | 16.0 (5.9) | 13.9 (3.4) | 18.5 (2) | .00<sup>B,C</sup> |
| tot gen | 18.3 (1.5) | 38.4 (8.8) | 31.7 (8.0) | 34.5 (2) | .00<sup>B,C</sup> |
| tot | 33.8 (2.4) | 73.7 (16.6) | 59.8 (14.9) | 38.4 (2) | .00<sup>B,C</sup> |
| QPE<sup>+</sup> |  |  |  | 20.1 (8,78) | .00 |
| severity VH | 0 (0) | 11.9 (4.4) | 0 (0) | 113.9 (2) | .00<sup>A,B,C</sup> |
| severity AH | 0 (0) | 13.1 (11.2) | 6.2 (9.7) | 9.2 (2) | .00<sup>B</sup> |
| severity all H | 0 (0) | 26.7 (13.2) | 6.8 (10.6) | 30.8 (2) | .00<sup>A,B</sup> |
| severity delusions | 0 (0) | 5.2 (5.8) | 4.6 (5.4) | 6.2 (2) | .00<sup>B,C</sup> |
| Use antipsychotics (n, (%)) |  |  |  |  |  |
| Yes |  | 10 (71.4) | 13 (86.7) | 1.0 (1) | .39 |
| No |  | 4 (28.6) | 2 (13.3) |  |  |
| Type antipsychotics (n, (%)) |  |  |  |  |  |
| 1 typical |  | 0 (0) | 1 (7.7) | 1.4 | .80 |
| 1 atypical |  | 6 (60.0) | 9 (69.2) |  |  |
| Multiple |  | 4 (40.0) | 3 (23.1) |  |  |
| Other medication<sup>l</sup> (n, (%)) |  |  |  | 0.21 | 1.00 |
| Yes, 1 | 3 (20.0) | 6 (42.9) | 6 (40.0) |  |  |
| No | 9 (60.0) | 5 (35.7) | 6 (40.0) |  |  |
| Multiple | 3 (20.0) | 3 (21.4) | 3 (20.0) |  |  |

PSVH+: patients with a psychotic disorder with visual hallucinations, PSVH-: patients with a psychotic disorder without visual hallucinations, HC: healthy controls.
<sup>A</sup> difference between PSVH+ and PSVH- ( $p < .05$ ), <sup>B</sup> difference between PSVH+ and HC ( $p < .05$ ), <sup>C</sup> difference between PSVH- and HC ( $p < .05$ ), \* 1: none, 2: elementary education, 8: university, <sup>#</sup> Mini-Mental State Examination<sup>18</sup>, <sup>^</sup> Digital Snellen chart, oculi dexter et sinister, <sup>~</sup> Contrast sensitivity (GECKO chart, maximum score 16), <sup>&</sup> Positive and Negative Syndrome Scale; total positive, negative and general symptom scores<sup>20</sup>, <sup>+</sup> Questionnaire for Psychotic Experiences, VH: visual hallucinations, AH: auditory hallucinations, H: hallucinations (QPE;
http://qpeinterview.com/home/<sup>19</sup>, <sup>1</sup> antidepressants: PSVH+ 7.1%, PSVH- 46.7%, HC 12.5%; mood stabilizer: PSVH+ 7.1%; benzodiazepines/anxiolytics: PSVH+ 14.3%, PSVH- 13.3%; stimulants: PSVH- 6.7%; akineton: PSVH+ 7.1%; other: PSVH+ 42.9%, PSVH- 13.3%, HC 37.5%.

The mean age of all participants was 34.7 years (SD 10.1); 71.1% (n=32) were male. The groups were matched for age, sex and cognitive profile (all participants had a MMSE score>25). Most patients were diagnosed with schizophrenia (55.2%), 20.7% were diagnosed with schizoaffective disorder, and 24.1% with psychotic disorder NOS. The mean disease duration of PSVH+ was 11.1 years (SD 9.0) and of PSVH-15.1 years (SD 12.6) (not significantly different). Patients had significantly higher scores compared to HC on all items assessing psychotic symptoms (PANSS, QPE). PSVH+ had the highest scores. PSVH+ scored significantly higher compared to PSVH-on the PANSS total positive symptoms and the QPE items on VH and the total hallucination score. Most patients used antipsychotics (PSVH+ 71.4%, PSVH-86.7%), which was usually an atypical antipsychotic (PSVH+ 60.0%, PSVH-69.2%).

Seven PSVH+ and three PSVH-experienced hallucinations during scanning. One PSVH+ saw ongoing flashes of lights, another occasionally saw people and spaceships, the third PSVH+ saw a dog or donkey 10 times for 1 second. Two of them also experienced nearly continuous auditory hallucinations (AH). One PSVH+ experienced nearly continuous AH and tactile hallucinations (TH). Two PSVH+ experienced only TH, one continuously and the other briefly. Another PSVH+ reported AH that lasted 2 minutes in total. One PSVH-participant experienced nearly continuous AH, and occasional TH. One PSVH-participant experienced a brief AH. finally, one PSVH-experienced continuous TH.

### Whole brain functional connectivity

Fig. 2 depicts whole brain FC matrices averaged per group and the FC distributions per group. Both PSVH+ and PSVH-had a lower overall FC than HC (HC versus PSVH+ p<0.041, HC versus PSVH-p<0.044). The overall FC scores of PSVH+ and PSVH-were very similar.

**Fig. 2).**
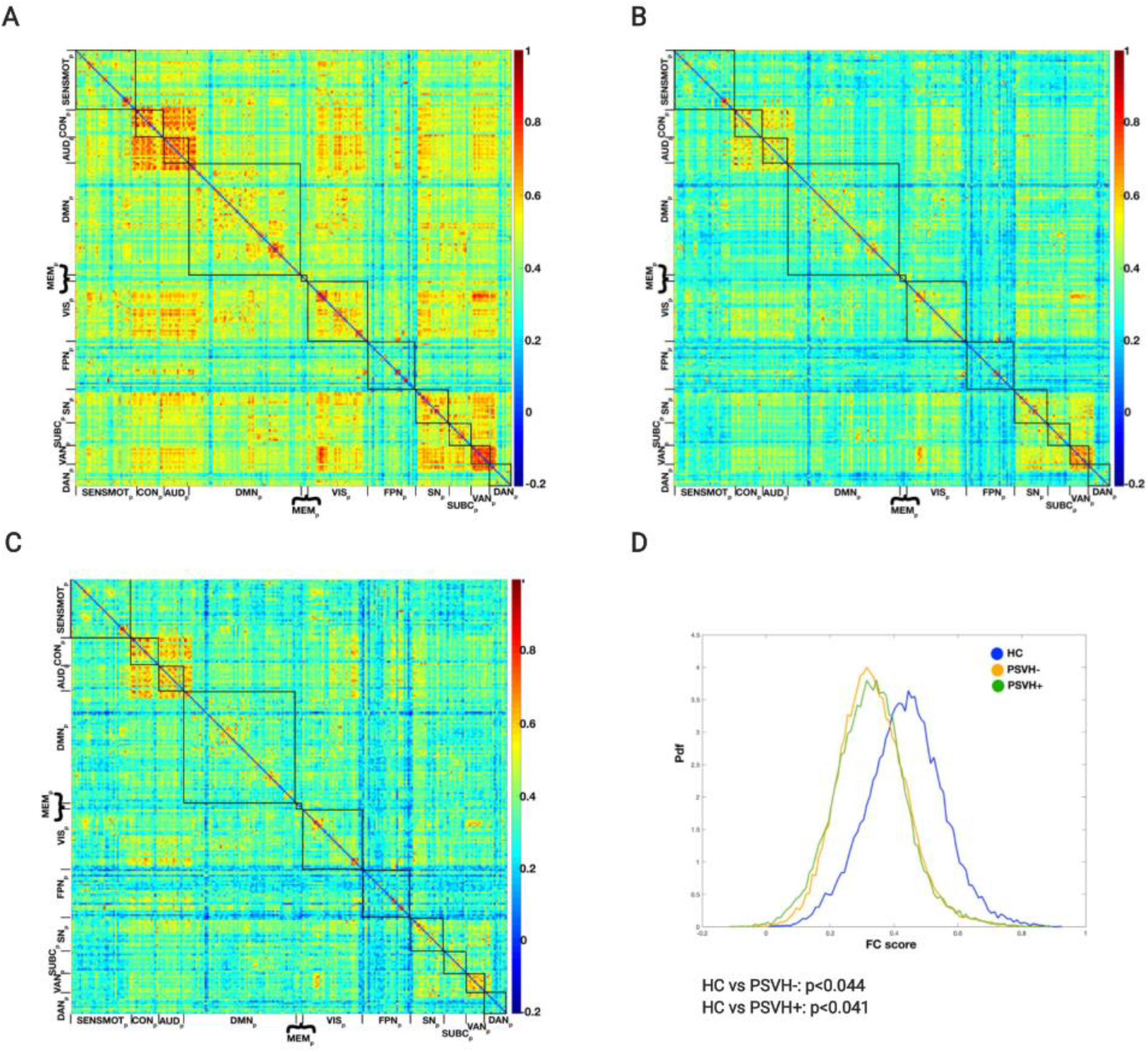
Whole brain functional connectivity. whole brain functional connectivity per group, based on the Power atlas^25^. SENSMOT_p_: Sensory/somatomotor Hand and Mouth Networks, CON_p_: Cingulo-opercular Task Control Network, AUD_p_: Auditory Network, DMN_p_: Default Mode Network, MEM_p_: Memory Retrieval Network, VIS_p_: Visual Network, FPN_p_: Fronto-parietal Task Control Network, SN_p_: Salience Network, SUBC_p_: Subcortical Network, VAN_p_: Ventral Attention Network, DAN_p_: Dorsal Attention Network. A: healthy controls (HC), B: patients with a psychotic disorder without visual hallucinations (PSVH-), C: patients with a psychotic disorder with visual hallucinations (PSVH+), D: functional connectivity distributions per group and significant (p≤ 0.05) group comparisons.

### Intra-network functional connectivity

Fig. 3 shows the intra-network FC for the six predefined networks (see Suppl. Fig. 2 for the results for the other networks. The median and 95% confidence interval values per group, covering all networks, are shown in Suppl. Table 1). PSVH+ had a lower intra-network FC than HC for VIS_p_ (p=.011), VAN_p_ (p=.01), SN_p_ (p=.016), DAN_p_ (p=.014) and DMN_p_ (p=.037). PSVH-had a lower intra-network FC compared to HC for only the DMN_p_ (p=.037). None of the networks showed a higher intra-network FC in PSVH+ and/or PSVH-compared to HC, and no significant differences between PSVH+ and PSVH-were found. To identify the most affected networks, Table 2 depicts their rank based on the ES when comparing the three groups. Notably, compared to both HC and PSVH-, the DAN and the VAN_p_ were affected most in PSVH+.

**Fig. 3).**
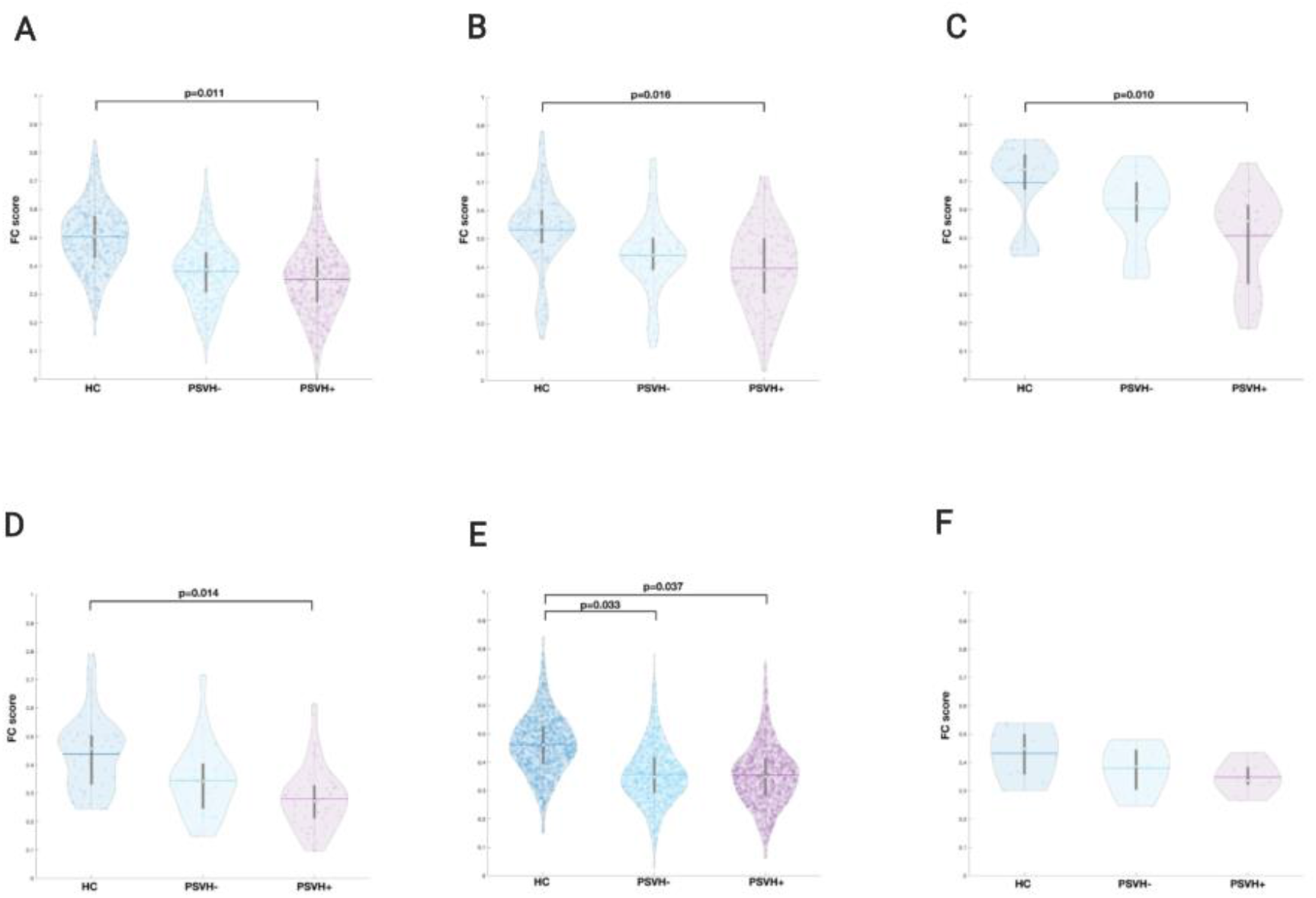
intra-network functional connectivity, per group and group comparisons. A: VIS_p_, Visual Network, B: VAN_p_, Ventral Attention Network, C: SN_p_, Salience Network, D: DAN_p_, Dorsal Attention Network, E: DMN_p_, Default Mode Network, F: MEM_p_, MEmory retrieval Network PSVH+: patients with a psychotic disorder with visual hallucinations, PSVH-: patients with a psychotic disorder without visual hallucinations, HC: healthy controls. Violin plots: white dot: median, horizontal stripe: mean.

**Table 2:** Ranking of most impaired intra-network functional connectivity based on effect size.

|  | HC vs PSVH+ | HC vs PSVH- | PSVH- vs PSVH+ |
| --- | --- | --- | --- |
| DAN<sub>p</sub> | 1 | 6 | 1 |
| VAN<sub>p</sub> | 2 | 5 | 2 |
| SN<sub>p</sub> | 3 | 3 | 3 |
| VIS<sub>p</sub> | 4 | 1 | 5 |
| MEM<sub>p</sub> | 5 | 4 | 4 |
| DMN<sub>p</sub> | 6 | 2 | 6 |

*Legend: PSVH+: patients with a psychotic disorder with visual hallucinations, PSVH-: patients with a psychotic disorder without visual hallucinations, HC: healthy controls.*
*VIS<sub>p</sub>*: Visual Network, *VAN<sub>p</sub>*: Ventral Attention Network, *SN<sub>p</sub>*: Salience Network, *DAN<sub>p</sub>*: Dorsal Attention Network, *DMN<sub>p</sub>*: Default Mode Network, *MEM<sub>p</sub>*: MEemory retrieval Network

### Inter-network functional connectivity

Fig. 4 shows the inter-network FC for the six predefined networks (see Suppl. Fig 3 for the other networks). PSVH+ showed a lower inter-network FC compared to HC for the following pairs: VIS_p_-SN_p_, VIS_p_-DAN_p_, VIS_p_-DMN_p_, SN_p_-VAN_p_, SN_p_-DAN_p_, SN_p_-DMN_p_, VAN_p_-DAN_p_, VAN_p_-DMN_p_, and the DAN_p_-DMN_p_. PSVH-had lower inter-network FC than HC for VIS_p_-DAN_p_, VIS_p_-DMN_p_, VAN_p_-DAN_p_, SN_p_-DAN_p_, VAN_p_-DMN_p_ and the DAN_p_-DMN_p_. No network pair showed a higher FC in PSVH+ than in PSVH- and/or HC. PSVH+ and PSVH-did not differ significantly for all network pairs. Table 3 depicts the ranking of the most affected network pairs, based on ES. Compared to both HC and PSVH-, the VAN_p_ and SN_p_ were the most affected networks in PSVH+.

**Fig. 4).**
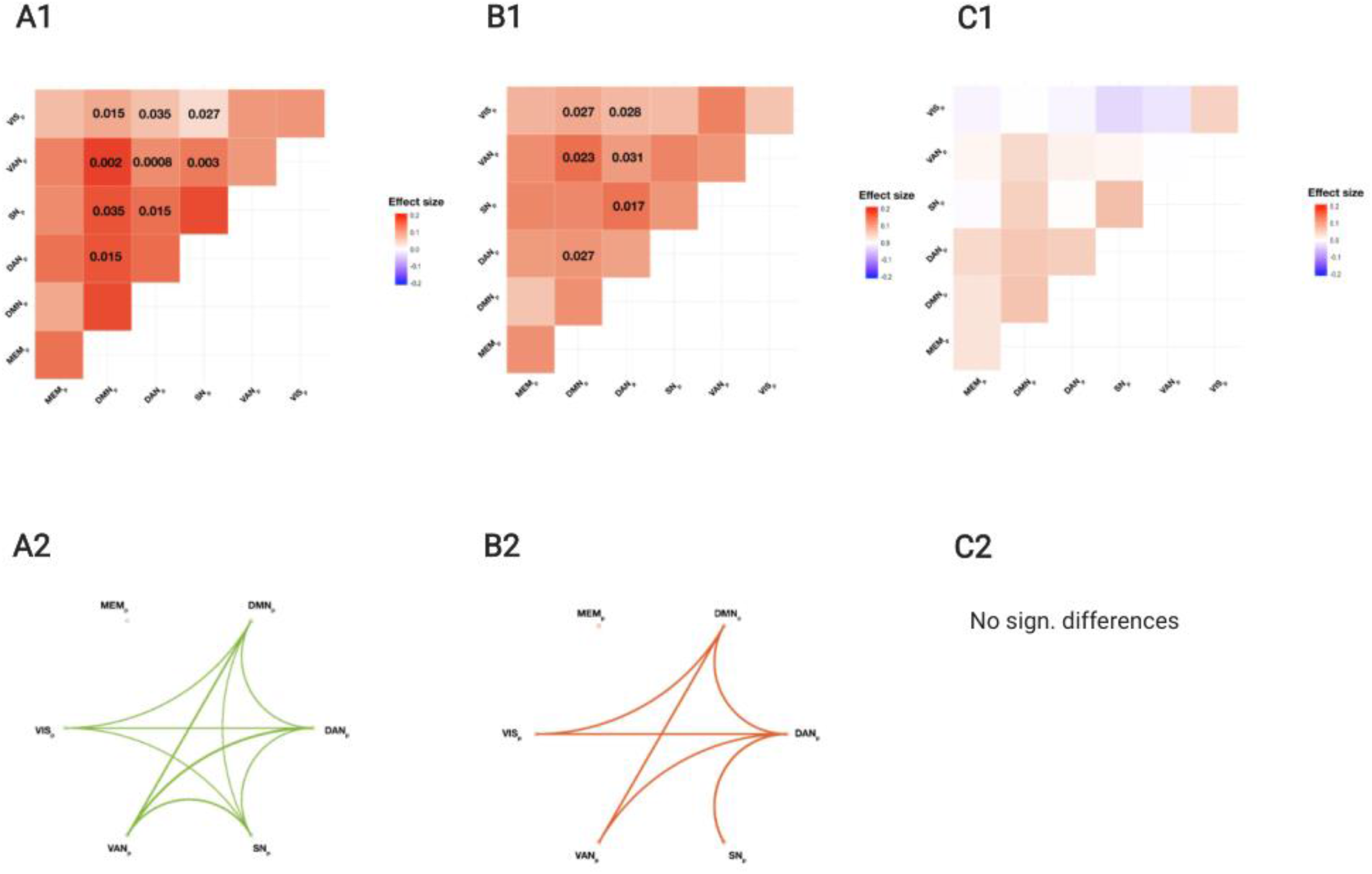
inter-network functional connectivity, group comparisons. A: healthy controls (HC) versus patients with a psychotic disorder with visual hallucinations (PSVH+), B: healthy controls (HC) versus patients with a psychotic disorder without visual hallucinations (PSVH-), C: patients with a psychotic disorder without visual hallucinations (PSVH-) versus patients with a psychotic disorder with visual hallucinations (PSVH+). 1: effect sizes of significant (p≤ 0.05) inter-network FC comparisons, 2: display of these significant differences. VIS_p_: Visual Network, VAN_p_: Ventral Attention Network, SN_p_: Salience Network, DAN_p_: Dorsal Attention Network, DMN_p_: Default Mode Network, MEM_p_: MEmory retrieval Network

**Table 3:** Ranking of most impaired inter-network functional connectivity based on effect size.

| NW pairs | Ranking |  |  |
| --- | --- | --- | --- |
|  | HC vs PSVH+ | HC vs PSVH- | PSVH- vs PSVH+ |
| VAN<sub>p</sub>-DMN<sub>p</sub> | 1 | 1 | 4 |
| VAN<sub>p</sub>-DAN<sub>p</sub> | 2 | 6 | 2 |
| **SN<sub>p</sub>-VAN<sub>p</sub>** | 3 | 8 | 1 |
| **SN<sub>p</sub>-DAN<sub>p</sub>** | 4 | 2 | 14 |
| **VIS<sub>p</sub>-SN<sub>p</sub>** | 5 | 9 | 3 |
| **VIS<sub>p</sub>-DMN<sub>p</sub>** | 6 | 7 | 10 |
| **VIS<sub>p</sub>-DAN<sub>p</sub>** | 7 | 4 | 12 |
| **DAN<sub>p</sub>-DMN<sub>p</sub>** | 7 | 4 | 12 |
| **SN<sub>p</sub>-DMN<sub>p</sub>** | 9 | 10 | 8 |
| **DMN<sub>p</sub>-MEM<sub>p</sub>** | 10 | 3 | 7 |
| **VIS<sub>p</sub>-VAN<sub>p</sub>** | 11 | 15 | 6 |
| **VAN<sub>p</sub>-MEM<sub>p</sub>** | 12 | 11 | 15 |
| **VIS<sub>p</sub>-MEM<sub>p</sub>** | 13 | 12 | 9 |
| **SN<sub>p</sub>-MEM<sub>p</sub>** | 14 | 13 | 11 |
| **DAN<sub>p</sub>-MEM<sub>p</sub>** | 15 | 14 | 5 |

*Legend: HC: healthy controls, PSVH+: patients with a psychotic disorder with visual hallucinations, PSVH-: patients with a psychotic disorder without visual hallucinations*
*VIS<sub>p</sub>: Visual Network, VAN<sub>p</sub>: Ventral Attention Network, SN<sub>p</sub>: Salience Network, DAN<sub>p</sub>: Dorsal Attention Network, DMN<sub>p</sub>: Default Mode Network, MEM<sub>p</sub>: MEemory retrieval Network*

### Eigenvector Centrality Mapping

Fig. 5 A-C displays the ROIs with the 5% highest centrality value per group. For all groups, this includes ROIs from the DMN_p_, VIS_p_ and VAN_p_/SN_p_. The ROIs with the 10% highest centrality value per group (Suppl. Fig. 1) belonged to the same networks. Back to the 5% highest centrality value, Fig. 5 D depicts the mean ECM values per group of the ROIs that differed significantly between groups. Three ROIs showed differences between PSVH+ and HC. These ROIs were the right intracalcarine cortex (ICalc, VIS), the left middle temporal gyrus temporo-occipitally (MTGo, VAN_p_) and for the lateral part of the left middle occipital gyrus (MOGl, VIS_p;_ almost reaching statistical significance). PSVH+ had lower ECM values for the right ICalc and left MTGo than HC, and a strong trend towards higher left MOGl values compared to HC. The ECM value of the right ICalc in PSVH+ is close to the null distribution. The ECM values were not significantly different between PSVH+ and PSVH- or between HC and PSVH-. Suppl. Fig 4. shows the mean ECM values of the ROIs within the highest 5 percentile, but these did not differ significantly between groups.

**Fig. 5).**
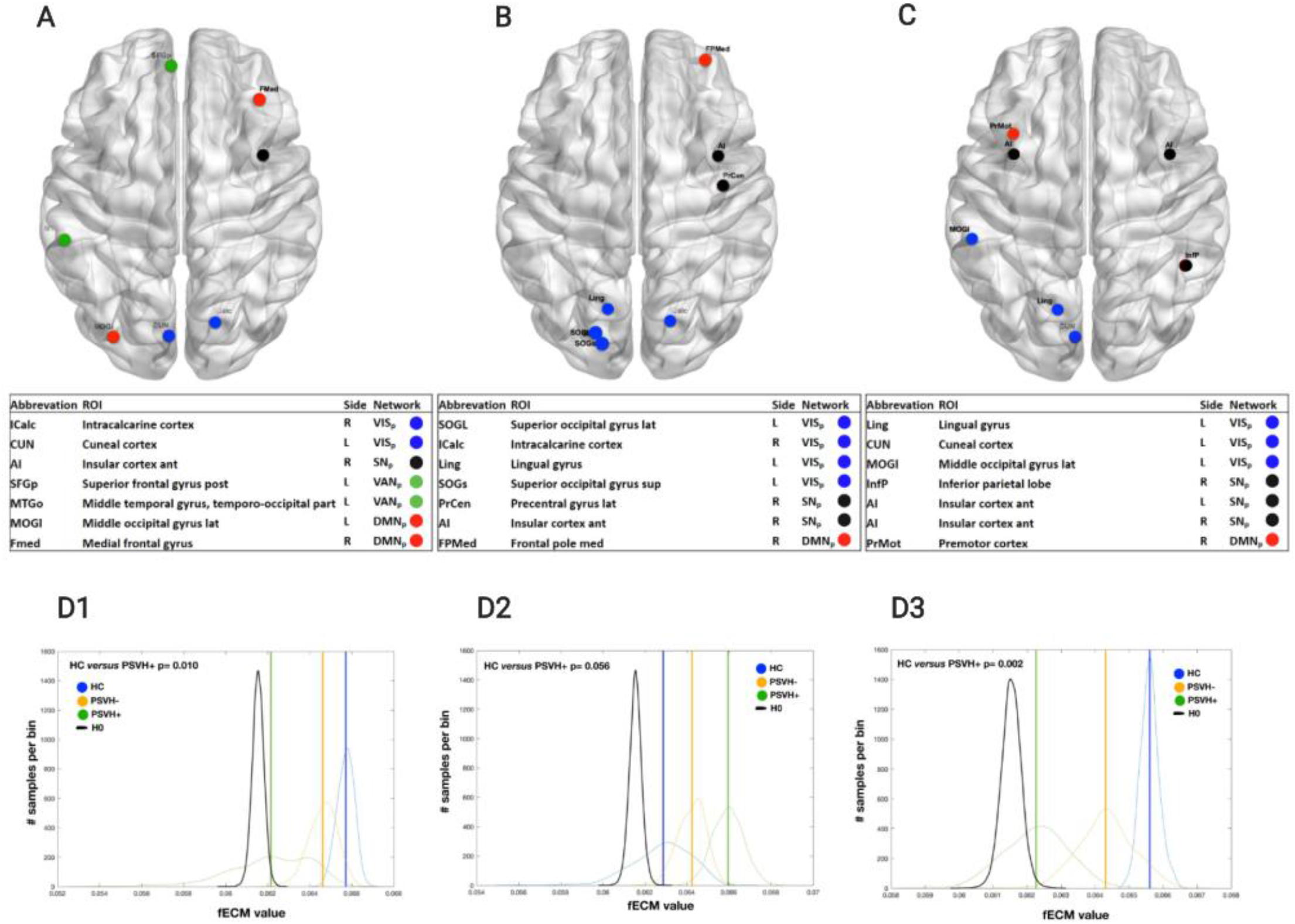
ROIs with 5% highest centrality value per group, and group differences. A: healthy controls (HC), B: patients with a psychotic disorder with visual hallucinations (PSVH+), C: patients with a psychotic disorder without visual hallucinations (PSVH-), D: mean ECM values per group of the ROIs that significantly (p≤ 0.05) differ between groups: 1: the right intracalcarine cortex (ICalc, VIS), 2: the left middle temporal gyrus temporo-occipital part (MTGo, VAN_p_) and 3: a trend for the left middle occipital gyrus lat (MOGl, VIS_p_; not significantly different). Note that the null distribution of the ECM is not centered at zero, as ECM values were positive real-valued. To define the confidence intervals of each ECM value estimated per ROI, a bootstrap technique (across time-points) was used at group level in parallel to resample the filtered fMRI data. To support visualization, a Gaussian distribution was fitted to both bootstrap and surrogate distributions.

## Discussion

Our study yielded three salient findings. First, we found that overall FC in patients is lower than in HC. Second, we found that intra- and inter-network FC is reduced in all the vision-related functional networks in PSVH+, the VAN being most affected. Third, in PSVH+ we found that the centrality is decreased in the right intracalcarine cortex and increased in the lateral part of the left middle occipital gyrus. Overall, our study supports the notion that the symptoms of schizophrenia do not result from focal pathology, but primarily from dysconnectivity between brain regions and networks ^3^. Below, we discuss these findings in more detail and the implications for understanding VH in psychosis. See Fig. 1 for an overview of the results (C1 and C2) together with an overview of vision-related networks (A) and the hypothesis (B).

### Lower whole brain functional connectivity in patients with psychosis

As hypothesized, PSVH+ and PSVH-had lower overall FC than HC ^4,32^. Our data confirms other rs-fMRI studies, which also found generally lower cerebral FC in schizophrenia than in controls ^4,33^. However, other studies reported higher FC in schizophrenia, involving the prefrontal cortex, inferior parietal lobe, medial regions, frontal and temporal lobe ^34^, cingulate gyrus and thalamus ^35^. This suggests a more complex picture of the generation of schizophrenia symptoms. In line with this, Venkataraman et al. ^36^ found coexisting FC patterns in schizophrenia: lower FC between parietal and temporal regions, and between the temporal cortices, related to positive symptoms, whereas higher FC between parietal and frontal regions was associated with negative and general symptoms.

### VH in psychosis relate to widespread lower intra-network and inter-network connectivity in vision-related networks

Based mainly on the model of Shine, we hypothesized that VHs in psychosis relate to lower intra-VIS, intra-VAN and intra-DMN FC, with higher inter-VIS-DMN FC ^1,10^. Our results only confirm part of this hypothesis, since we found lower inter-VIS_p_-DMN_p_ FC. Moreover, our results indicate that widespread FC impairments in vision-related networks is associated with the occurrence of psychosis-related VH. Whereas PSVH-only showed lower intra-DMN_p_ FC compared to HC, PSVH+ showed a decrease in almost all the vision-related networks (VIS_p_, VAN_p_, SN_p_, DAN_p_ and DMN_p_). Similarly, PSVH-showed reduced inter-network FC for various network pairs compared to HC. However, PSVH+ showed even more reductions, mainly involving the VAN. Generally, PSVH-FC values were in between PSVH+ and HC (although not statistically tested). Our results correspond with multiple studies in schizophrenia that have shown widespread altered FC. A recent meta-analysis reported reduced intra-network FC for the DMN, affective network, VAN, thalamus network and the somatosensory network in schizophrenia ^13^. This study also found lower inter-network FC between VAN-TN, VAN-DMN, VAN-FrontoParietal central executive Network (FPN, see below), FPN-thalamus network, FPN-DMN, and higher FC for VAN-affective network in schizophrenia. Accordingly, early-stage schizophrenia was related to reduced inter-network FC between the somatomotor-limbic system, somatomotor system-DMN, DAN-DMN, VAN-limbic system, and VAN-DMN ^37^.

Wang et al. more specifically identified the intra-network and inter-network FC between large-scale networks belonging to Menon’s ‘triple-network model’ for positive symptoms in schizophrenia ^38–40^. It includes the SN (which corresponds to the VAN of Shine et al. VAN ^1^), DMN, and the FPN ^39^. Similar to the DAN, the FPN is a task-positive network. It includes the dorsolateral prefrontal and posterior parietal cortices, involving goal-directed higher order cognition such as attention, working memory and decision-making ^41^. Deficits in the engagement and disengagement of these three core neurocognitive networks play a key role in schizophrenia ^38^. Accordingly, patients with schizophrenia had a reduced inter-network FC for SN-DMN, SN-FPN and FPN-DMN ^39^. An important aspect of this model is the inappropriate assignment of saliency to external and internal stimuli by the SN ^38^. Aberrant SN activity causes dysfunctional switching between the DMN and FPN. Accordingly, intra-SN FC was reduced ^39^. Overall, our results corroborate the models proposed by both Shine et al. and Menon ^1,38^. Note that the model of Shine et al. in schizophrenia was partially based on auditory hallucinations, while Menon’s model was developed for positive symptoms in schizophrenia. In in our study, however, we focused on VH: PSVH+ and PSVH-participants showed a high prevalence of auditory hallucinations. Therefore, an auditory component in our results cannot be ruled out completely.

As most patients with psychosis with VH also have auditory hallucinations ^42^, it is difficult to disentangle their mechanisms. This may suggest a partially shared underlying mechanism.

### The Ventral Attentional Network and its role in psychosis-related VH

Both the model proposed by Shine et al. ^1^ and the triple brain network model in schizophrenia by Menon indicate that the VAN is most affected in VH and schizophrenia ^38^. Our inter-network analysis in PSVH+ supports the notion that the VAN_p_ is the most impaired network Moreover, our intra-network analysis indicated that it was the second most impaired network, and Eigenvector analyses indicated that its left MTGo showed less communication compared to HC. The MTGo is involved in saliency processing by detecting changes in the visual environment in color, shape, orientation and motion ^43–45^. PSVH+ showed the highest impairment of intra-network FC in the DAN_p_. A previous study also found lower intra-DAN FC during rs-fMRI compared to controls ^46^. However, other studies found higher intra-DAN FC and higher DAN-activity during a target detection task in schizophrenia ^47,48^.

Menon et al. suggested that suboptimal functioning of the VAN (which includes the SN), together with inadequate switching between the DMN and DAN, could lead to impaired salience mapping and, consequently, to confusion between stimuli from the internal and external world, thus causing VH ^38^. Moreover, the DAN’s inability to appropriately prime salient phenomena would contribute to the impaired interpretation of visual stimuli ^1^. Although our study did not address causality, our findings appear to support these models. Interestingly, multiple studies point towards a SN dysfunction as the root of the impaired inter-network FC. In schizophrenia, intra-SN variation negatively predicts the FC between the three networks, which is mediated by intra-SN connectivity ^39^. Moreover, in schizophrenia the SN-FPN and SN-DMN interactions were also weaker, and the switches between these networks were more frequent ^40^. These alterations correlated positively with positive symptoms, including hallucinations. Graph theory and causal reasoning also showed that the independent components with the most influential roles in producing auditory hallucinations-related activity were those within the SN ^49^.

### VH in psychosis are related to impaired functional communication within the occipital cortex

Corresponding with the model of Shine et al., we found that the intra-VIS FC in PSVH+ was reduced. This indicates that VHs in psychosis are related to impaired communication between visual areas additional to widespread impairments described above. Furthermore, in our study the ECM value for the right ICalc was close to the null distribution in PSVH+. At the same time, the left MOGl in PSVH+ was more central compared to HC. This change in centrality can be interpreted as a decrease in information transfer (communication) by the ICalc and an increase by the MOGl. Note that the ICalc definition used in our study includes V1 ^50^, while the MOGl is part of the lateral occipital complex (LOC), a higher visual area, mainly involved with processing of faces ^51^, animals ^52^ and objects ^53^. Based on our data, we conclude that VH are related to severely reduced communication between V1 and other brain areas, and at the same time to increased communication between the LOC and other areas, except V1. Other studies also reported occipital functional segregation in schizophrenia ^4,5^. Because PSVH-did not differentiate from HC, our study suggests that this higher occipital segregation is more specifically related to VH, instead of being related to schizophrenia in general. This suggests that VH in psychotic patients are related to deficits in the early stages of visual processing, which very likely contributes to the impaired salience detection by the VAN. The increased central role of the LOC corresponds with the predominantly complex nature of VH in psychosis ^7,8,54,55^.

Our study has several limitations. Firstly, multiple (3) participants reported VH during scanning, which may have influenced our results. For example, during VH in psychosis the DMN showed lower activity than during periods without VH ^56^. Secondly, medication may have affected our results. For ethical reasons, participants were not taken off their medication. However, antipsychotics influence dopaminergic signaling, whereas dopamine plays an important role in coupling the explored networks ^57^. Thirdly, our study did not include causal modeling.

We would recommend future studies to use causal methodologies to determine whether a similar causal mechanism as for auditory hallucinations (with a key role for the VAN) underlies VH in psychosis. Noteworthy, this study only addressed six networks that are directly related to vision.

Nonetheless, schizophrenia is known to be related to widespread dysconnectivity (see section ‘VH in psychosis relate to widespread lower intra-network and inter-network connectivity in vision-related networks’ in the discussion). The results for the other networks that are not directly related to vision are presented in Suppl. Fig. 2 and 3. These results may lead to new research questions. For example, we unexpectedly found that PSVH+ have lower intra-NW FC of the subcortical areas compared to HC, and lower inter-network FC between subcortical areas and the VAN compared to PSVH-. Last, this study included participants between 18-55 years old who mostly lived in the northern part of the Netherlands. Future studies will show if our results also hold true for other populations.

## Conclusion

We found that widespread dysconnectivity of predominantly vision-related functional networks predisposes patients with psychosis to generate visual hallucinations, with this decrease being most prominent for the Ventral Attention Network, and for the Dorsal Attention Network for intra-network functional connectivity. Our results are therefore in line with earlier models on hallucinations in psychosis stating that the processing deficits in the Visual Network may either result in or exacerbate inadequate co-functioning and switching between the Default Mode Network and Dorsal Attention Network, possibly due to impaired Ventral Attention Network functioning. This, in combination with impaired attending of visual signals by the DAN, may lead to inappropriate saliency processing and wrongly attributing an external origin to internally generated events and, consequently, to VH. The mostly complex nature of the psychotic visual hallucinations may be explained by the more central role assumed by the LOC in visual processing, as observed during resting state activity.

## Supporting information

Suppl. Fig. 1 fECM 90

Suppl. Fig. 2 intra-network FC other networks

Suppl. Fig. 3 inter-network FC all networks

Suppl. Fig. 4 fECM non-sign ROIs

Suppl. Table 1 intra-network FC median and 95%CI

strobe

## Acknowledgements

We would like to thank Wendy Groot Jebbink, Martijn Majoor, Wouter Staal, Anita Sibeijn-Kuiper for their help in fMRI data acquisition.

## Supplementary Material

***Legends***

***Suppl. Fig. 1***

*A: healthy controls (HC), B: patients with a psychotic disorder with visual hallucinations (PSVH+), C: patients with a psychotic disorder without visual hallucinations (PSVH-)*

***Suppl. Fig. 2***

*A: Sensory/somatomotor Hand and Mouth Networks (SENSMOT*_*p*_*), B: Cingulo-opercular Task Control Network (CON*_*p*_*), C: Auditory Network (AUD*_*p*_*), D: Fronto-parietal Task Control Network (FPN*_*p*_*), E: Subcortical Network (SUBC*_*p*_).

*HC: healthy controls, PSVH+: patients with a psychotic disorder with visual hallucinations, PSVH-: patients with a psychotic disorder without visual hallucinations*

*Violin plots: white dot: median, horizontal stripe: mean*.

***Suppl. Fig. 3***

*SENSMOT*_*p*_: *Sensory/somatomotor Hand and Mouth Networks, CON*_*p*_: *Cingulo-opercular Task Control Network, AUD*_*p*_: *Auditory Network, DMN*_*p*_: *Default Mode Network, MEM*_*p*_: *Memory Retrieval Network, VIS*_*p*_: *Visual Network, FPN*_*p*_: *Fronto-parietal Task Control Network, SN*_*p*_: *Salience Network, SUBC*_*p*_: *Subcortical Network, VAN*_*p*_: *Ventral Attention Network, DAN*_*p*_: *Dorsal Attention Network*.

*HC: healthy controls, PSVH+: patients with a psychotic disorder with visual hallucinations, PSVH-: patients with a psychotic disorder without visual hallucinations*

***Suppl. Fig. 4***

*Mean ECM values per group of the ROIs that do not significantly differ between groups. Note that the null distribution of the ECM is not centered at zero, as ECM values were positive real-valued. To define the confidence intervals of each ECM value estimated per ROI, a bootstrap technique (across time-point) was used at group level in parallel to resample the filtered fMRI data. To support visualization, a Gaussian distribution was fitted to both bootstrap and surrogate distributions*.

*A: cuneal cortex (CUN) l, VIS*_*p*_; *B: lingual gyrus (Ling) l, VIS*_*p*_; *C: superior occipital gyrus sup (SOGs) l, VIS*_*p*_; *D: superior occipital gyrus lat (SOGl) l, VIS*_*p*_; *E: middle occipital gyrus lat (MOGl) l, VIS*_*p*_ ; *F: superior frontal gyrus post (SFGp) l, VAN*_*p*_; *G: precentral gyrus lat (PrCen) r, SN*_*p*_; *H: insular cortex ant (AI) r, SN*_*p*_; *I: insular cortex ant (AI) l, SN*_*p*_; *J: medial frontal gyrus (FMed) r, DMN*_*p*_; *K: premotor cortex (PrMot) r, DMN*_*p*_; *L: frontal pole med (FPMed) r, DMN*_*p*_.

*HC: healthy controls, PSVH+: patients with a psychotic disorder with visual hallucinations, PSVH-: patients with a psychotic disorder without visual hallucinations*

