## Supplementary figures and images for "Impaired functional connectivity in patients with psychosis and visual hallucinations"

### Suppl. Fig. 1 fECM 90

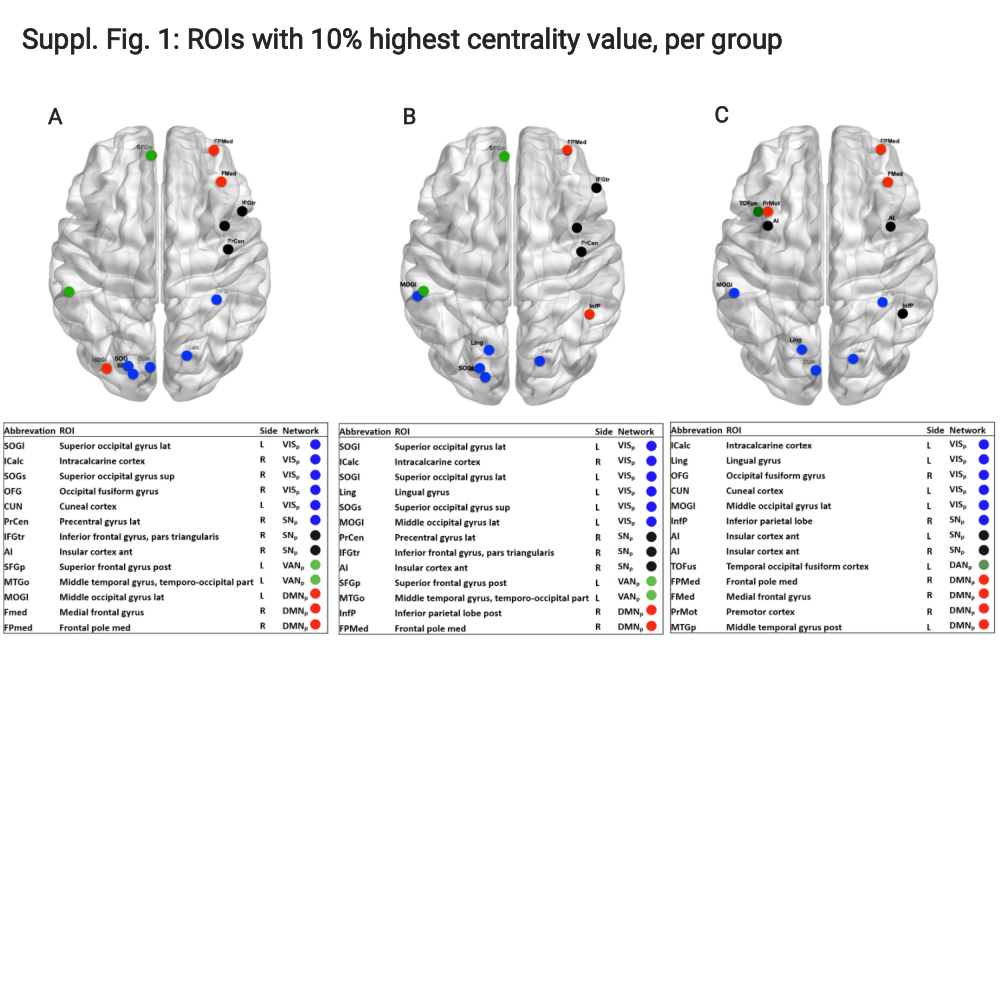

### Suppl. Fig. 2 intra-network FC other networks

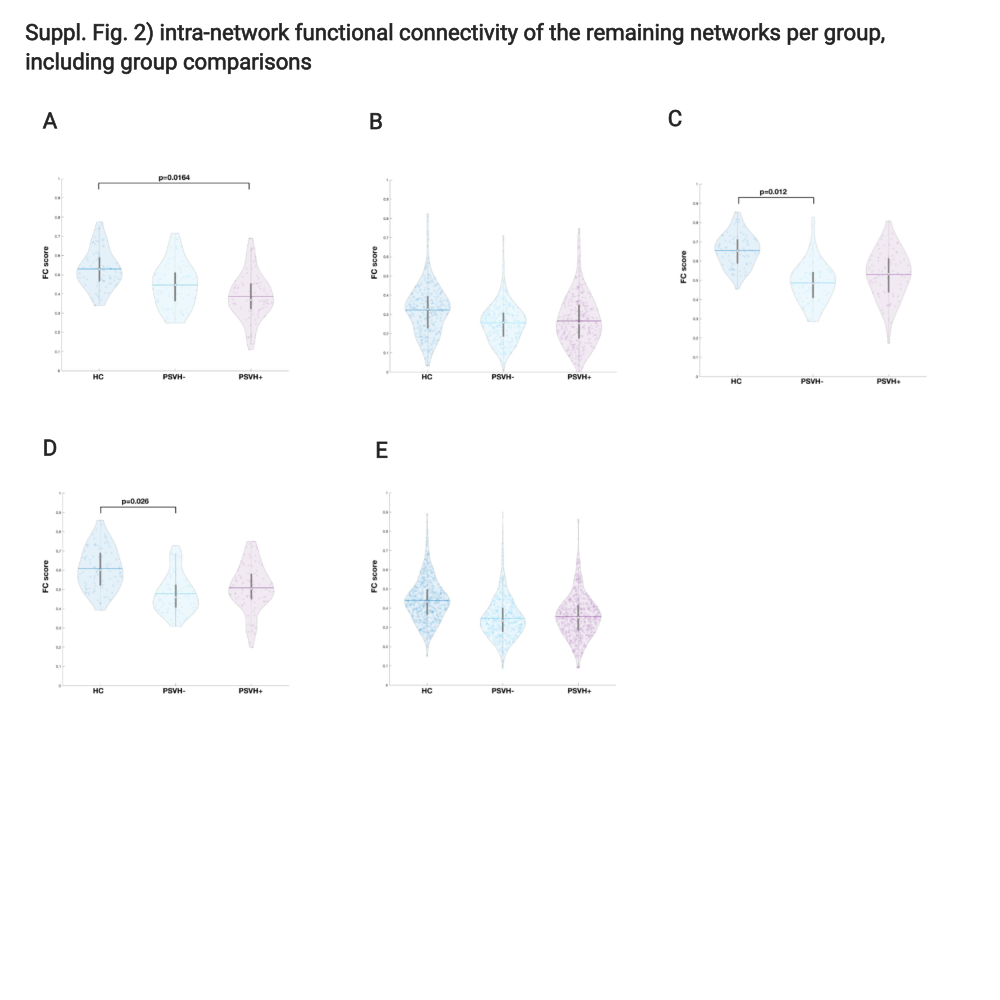

### Suppl. Fig. 3 inter-network FC all networks

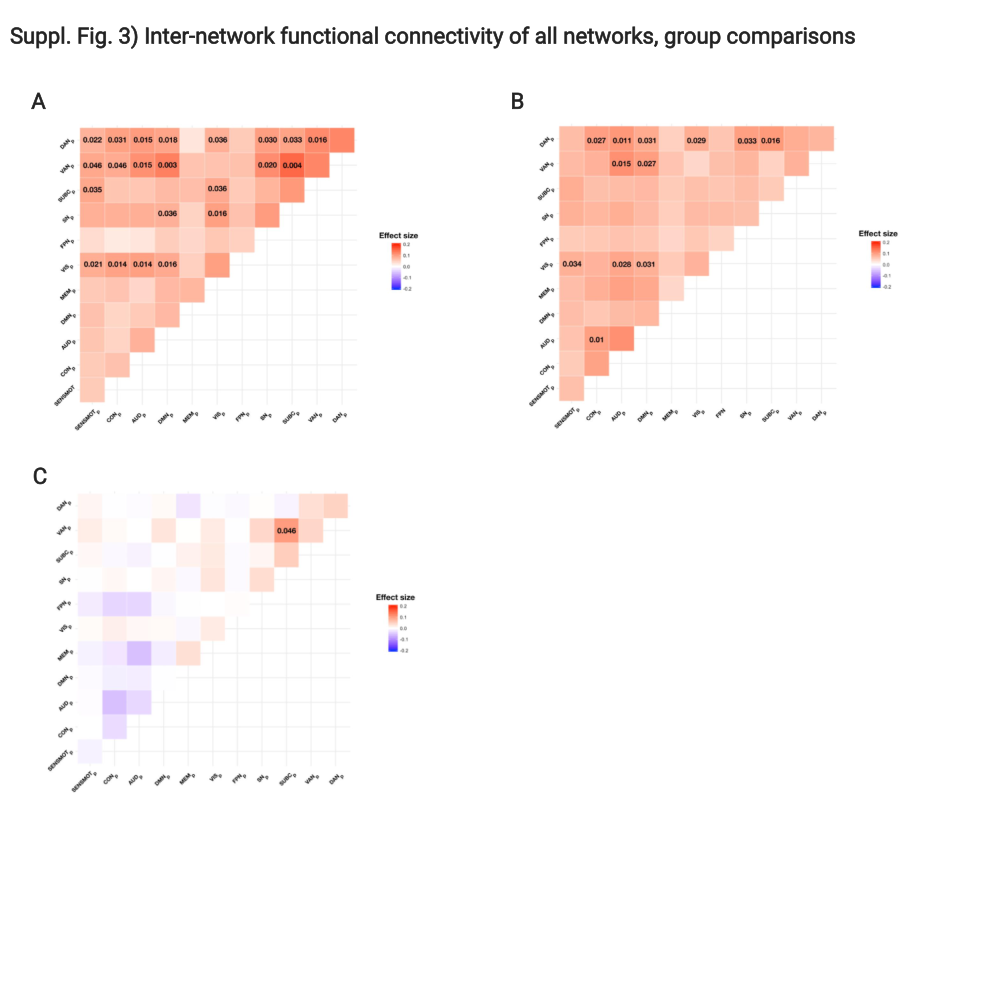

### Suppl. Fig. 4 fECM non-sign ROIs

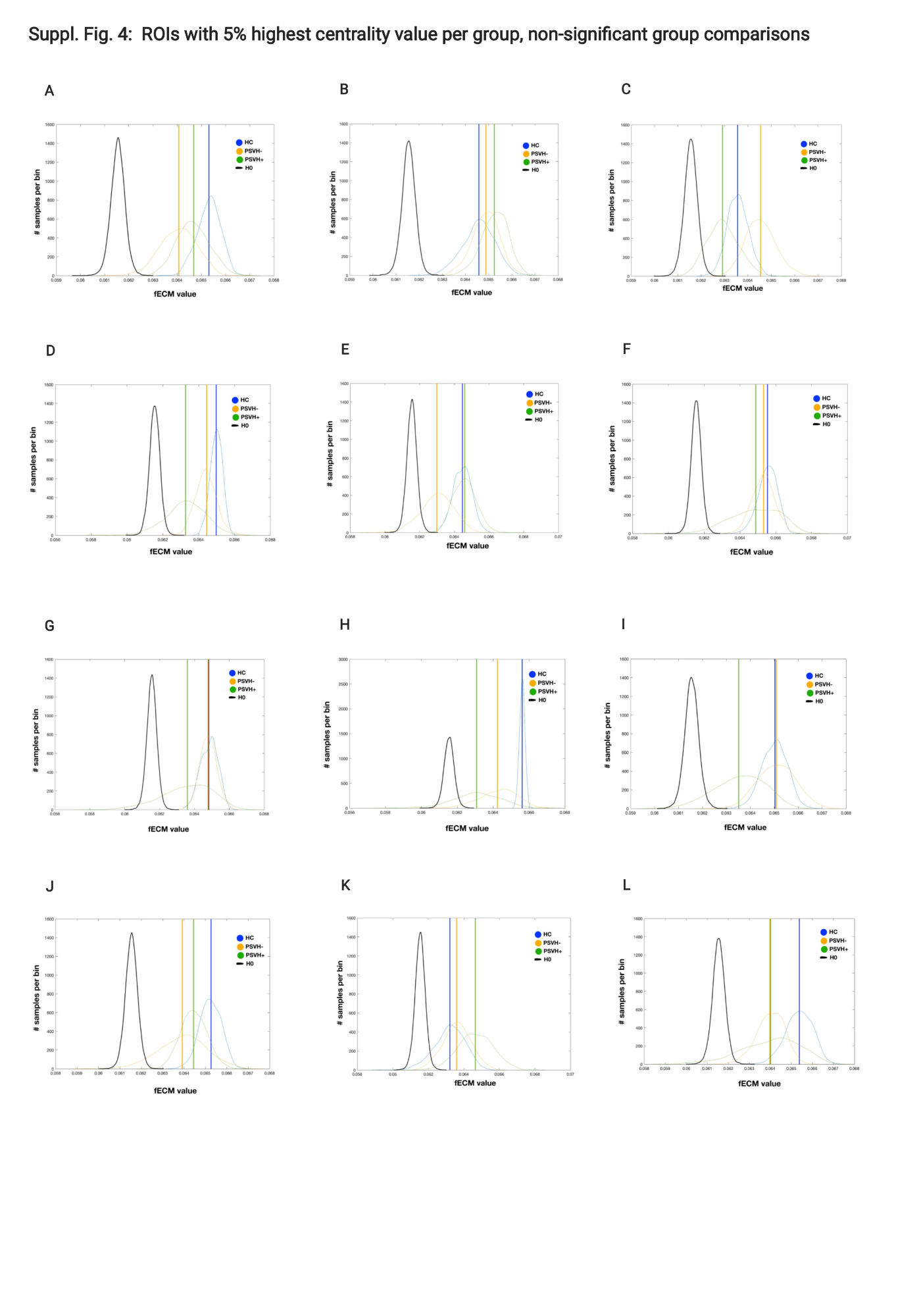
