## Supplementary material for "Impaired functional connectivity in patients with psychosis and visual hallucinations": Suppl. Table 1 intra-network FC median and 95%CI

**Supplementary Table 1: intra-network functional connectivity: median and 95% confidence-interval per group**

| Network | Median [95% CI] | | |
|---|---|---|---|
| | HC | PVSH- | PVSH+ |
| $VIS_p$ | 0.5039 [0.276 - 0.755] | 0.3857 [0.172 - 0.618] | 0.3549 [0.115 - 0.647] |
| $VAN_p$ | 0.7416 [0.436 - 0.845] | 0.6221 [0.357 - 0.192] | 0.5585 [0.192 - 0.742] |
| $SN_p$ | 0.5432 [0.204 - 0.806] | 0.4431 [0.151 - 0.733] | 0.3898 [0.126 - 0.680] |
| $DAN_p$ | 0.4559 [0.245 - 0.746] | 0.3422 [0.148 - 0.705] | 0.2723 [0.097 - 0.582] |
| $DMN_p$ | 0.4604 [0.263 - 0.688] | 0.3478 [0.159 - 0.593] | 0.3490 [0.158 - 0.461] |
| $MEM_p$ | 0.4481 [0.299 - 0.541] | 0.3850 [0.247 - 0.481] | 0.3359 [0.265 - 0.436] |
| $SENSMOT_p$ | 0.4335 [0.255 - 0.683] | 0.3341 [0.177 - 0.607] | 0.3514 [0.163 - 0.612] |
| $CON_p$ | 0.6008 [0.428 - 0.816] | 0.4588 [0.327 - 0.685] | 0.5039 [0.260 - 0.728] |
| $AUD_p$ | 0.6534 [0.523 - 0.816] | 0.4832 [0.289 - 0.744] | 0.5329 [0.288 - 0.738] |
| $FPN_p$ | 0.3262 [0.103 - 0.593] | 0.2569 [0.094 - 0.480] | 0.2528 [0.057 - 0.583] |
| $SUBC_p$ | 0.5274 [0.351 - 0.751] | 0.4457 [0.263 - 0.685] | 0.3683 [0.154 - 0.637] |
